# Examining neurophysiological markers of apathy and processing speed in late premanifest and early-stage manifest Huntington’s disease

**DOI:** 10.1101/2022.08.09.22278610

**Authors:** Marie-Claire Davis, Aron T. Hill, Paul B. Fitzgerald, Neil W. Bailey, Julie C. Stout, Kate E. Hoy

## Abstract

**Objective:** To find sensitive biological markers of non-motor symptoms in Huntington’s disease (HD), which are essential for the development and assessment of novel treatments.

**Methods:** We used resting state EEG to examine differences in oscillatory activity (analysing the isolated periodic as well as the complete EEG signal) and functional connectivity in 22 late premanifest and early stage people with HD and 20 neurotypical controls. We then assessed the correlations between these neurophysiological markers and clinical measures of apathy and cognitive functioning.

**Results:** Significantly lower theta and greater delta resting state power was seen in the HD group, as well as significantly greater delta connectivity. There was a significant positive correlation between theta power and processing speed, however there were no associations between the neurophysiological and apathy measures.

**Conclusions:** We speculate that these changes in oscillatory power and connectivity reflect ongoing, frontally concentrated degenerative and compensatory processes associated with HD.

**Significance:** Our findings support the potential utility of quantitative EEG as a proximate marker for non-motor symptoms in HD.

**Highlights:**

- We examined EEG oscillatory power and functional connectivity as a marker of non-motor symptoms in HD.
- The HD group had lower theta power, higher delta power and connectivity, and theta power was correlated with processing speed.
- Our findings support the use of quantitative EEG metrics as potential non-motor symptom markers for clinical research in HD.

## 1.1 Introduction

Huntington’s disease (HD) is an autosomal dominant trinucleotide repeat (cytosine adenine guanine; CAG) genetic disorder affecting the HTT gene, which results in a cascade of cellular, gross structural and functional neurophysiological changes (Pender & Koroshetz, 2011; Podvin, Reardon, Yin, Mosier, & Hook, 2019; Tyebji & Hannan, 2017; Waldvogel, Kim, Tippett, Vonsattel, & Faull, 2015). The currently accepted diagnostic criteria for ‘manifest’ or ‘symptomatic’ HD requires the presence of motor signs rated by an experienced clinician (preferably a neurologist) using the Total Motor Scale of the Unified Huntington’s Disease Rating Scale (UHDRS®) (Kieburtz, 1996; Oosterloo et al., 2021; Tabrizi et al., 2009). There is also extensive evidence of a prodromal disease stage characterised by cognitive and psychiatric changes, as well as subtle motor signs in the years prior to obvious motor symptom onset (Ghosh & Tabrizi, 2018; Paulsen, Miller, Hayes, & Shaw, 2017; Ross et al., 2019). This prodromal or late premanifest stage has recently been suggested for inclusion in the formal diagnostic criteria (Ross et al., 2019). The most common psychiatric symptoms during the late premanifest and early manifest stages of HD include anxiety, depression, irritability, and apathy (Andrews et al., 2020; Atkins, Andrews, Stout, & Chong, 2020; Epping et al., 2016; Martinez-Horta et al., 2016; Van Duijn et al., 2014). Apathy refers to “a lack of motivation that persists over time and causes identifiable functional impairment” (Robert et al., 2018, p. 72). Apathy is highly prevalent from the late premanifest through to advanced stages of HD and worsens with disease progression (Martinez-Horta et al., 2016; Paoli et al., 2017; Paulsen et al., 2017; Thompson et al., 2012). Mild cognitive impairment characterised by deficits in social cognition, attentional control, executive functions, and processing speed are also typically detected in neuropsychological examinations during the late premanifest and early manifest stages of HD (Duff et al., 2010; Langley et al., 2021; Paulsen et al., 2017; Stout, Andrews, & Glikmann-Johnston, 2017; Stout et al., 2014; Tabrizi et al., 2013). Reduced processing speed in particular is the earliest cognitive deficit to emerge in HD and linearly declines with disease progression (Paulsen et al., 2017).

Apathy and cognitive deficits such as reduced processing speed are debilitating for those in the late premanifest and early stages of manifest HD, impacting their ability to participate in home life, work and social activities (Beglinger et al., 2010; Eddy & Rickards, 2015; Fritz et al., 2018; Hergert & Cimino, 2021; Jacobs, Hart, & Roos, 2018; Vamos, Hambridge, Edwards, & Conaghan, 2007; Van Der Zwaan, Jacobs, van Zwet, Roos, & De Bot, 2021). Despite the functional impact of these symptoms, very few therapeutic interventions have been trialled, and to date, none have proven clinically effective (Dash & Mestre, 2020; Ferreira et al., 2022; Gelderblom et al., 2017).

In the context of resting state EEG, spectral power refers to amplitude (squared) of macroscopic neuroelectric activity, while functional connectivity refers to synchronisation between spatially discrete electrodes or regions of interest (Cohen, 2014; Friston, 1994). There is extensive evidence of the association between these neurophysiological indices and various aspects of cognition, emotion, and behaviour in diverse populations (Başar & Güntekin, 2013; Bertrand et al., 2016; Choi et al., 2021; Herrmann, Strüber, Helfrich, & Engel, 2016; Iyer et al., 2015; Klimesch, 1999; Knyazev, 2012; Nobukawa et al., 2020; Sanchez-Dinorin et al., 2021). In HD, altered spectral power in the delta and alpha frequency bands, and increased functional connectivity in the delta band have been consistently identified (Delussi, Nazzaro, Ricci, & De Tommaso, 2020; Hunter, Bordelon, Cook, & Leuchter, 2010; Odish, Johnsen, van Someren, Roos, & van Dijk, 2018; Painold et al., 2010; Piano, Imperatori, et al., 2017; Piano, Mazzucchi, et al., 2017; Ponomareva et al., 2014). Altered spectral power and connectivity have also been found to correlate with performance on clinical indices and cognitive tests (including measures of processing speed) in people with HD; including the UHDRS® total motor scale, number of CAG repeats on the affected allele, and performance on the Stroop Word Reading Test, Mini Mental Status Examination, verbal fluency (FAS) and the Symbol Digit Modalities Test (Delussi et al., 2020; Hunter et al., 2010; Odish et al., 2018; Painold et al., 2010; Piano, Imperatori, et al., 2017; Piano, Mazzucchi, et al., 2017; Ponomareva et al., 2014). Whether these neurophysiological markers relate to apathy in HD, however, has not been investigated.

Previous studies of resting state EEG in HD have also analysed narrowband spectral power across different frequency ranges without accounting for broadband non-oscillatory (aperiodic) activity which follows a 1/f distribution and contributes significant power to the EEG signal, particularly in lower frequencies (Donoghue et al., 2020; Kosciessa, Grandy, Garrett, & Werkle-Bergner, 2020). As such, it is not clear whether the elevated delta power in HD is produced by differences in delta oscillations specifically, or differences in the 1/f aperiodic activity. Recently, methods have become available for differentiating between the periodic and aperiodic components of the signal; an analysis approach that can confirm true differences in neural oscillatory amplitude by accounting for the 1/f aperiodic activity (Donoghue et al., 2020; Kosciessa et al., 2020). The 1/f aperiodic activity itself has also been associated with cognition and clinical conditions, so assessing whether it differs in HD is valuable for enhancing our understanding of this disorder (Peterson, Rosen, Belger, Voytek, & Campbell, 2021). A clearer understanding of spectral power and functional connectivity and their relationship with non-motor symptoms will assist with treatment development in HD.

Sensitive biological markers of non-motor symptoms in HD are essential for the development and assessment of novel treatments. Neurophysiological markers may have particular potential in this space. To this end we used resting state EEG to examine differences in the complete EEG signal, as well as isolated periodic activity and functional connectivity, in 22 people with HD and 20 neurotypical controls. We examined the correlations between these neurophysiological markers and clinical measures of apathy and cognitive functioning (specifically processing speed). We hypothesised that previously identified differences in resting state power and functional connectivity between people with HD and neurotypical controls would be replicated, and these neurophysiological differences would correlate with clinical measures of apathy and processing speed in people with HD.

## 2.1 Methods

### 2.2 Participants

We recruited 20 neurotypical controls and 22 people with HD aged between 18 and 75 years as part of randomised single session within subjects’ study of non-invasive brain stimulation for apathy in HD conducted between February 2019 and August 2021 (Australian New Zealand Clinical Trials Registry (ANZCTR): 12619000870156). This paper presents the results of the analysis of baseline resting state EEG data and sample characteristics. The results of the non-invasive brain stimulation investigation will be presented in separate papers. All participants in the HD group had genetically confirmed HD and were in the late premanifest (n = 11) or early manifest stage 1 (n = 11) of their disease, with a disease burden score (DBS) of ≥280 (DBS = [CAG repeats - 35.5] x current age), and all had a total functional capacity score of 9 or higher (maximum score of 13) (Ghosh & Tabrizi, 2018; Kieburtz, 1996; Langbehn, Brinkman, Falush, Paulsen, & Hayden, 2004; Penney, Vonsattel, MacDonald, Gusella, & Myers, 1997). The TFC assesses how much assistance a person with HD requires to perform tasks in five functional domains that decline with disease progression (i.e., occupation, finances, domestic chores, activities of daily living, and care level) (Beglinger et al., 2010). A score of ≥7 corresponds with the early stages (i.e., stages I and II), during which the person with HD has motor symptoms, but is able to manage their domestic responsibilities and perform their usual activities of daily living (Ghosh & Tabrizi, 2018; Shoulson & Fahn, 1979). During stage II the person may have had to reduce their level of occupational engagement (e.g., work hours or responsibilities) and may require some assistance with managing their financial affairs (Ghosh & Tabrizi, 2018; Shoulson & Fahn, 1979). We chose to recruit people with HD in the late premanifest and early manifest stages of the disease continuum, with no or only mild cognitive and/or motor symptoms, but enough variability in clinical presentation to enable correlational analyses. We also chose to recruit people in the late premanifest and early manifest stages because of the prevalence and functional impact of apathy and mild cognitive impairment on their continued participation in home, work, and social activities (Ghosh & Tabrizi, 2018; Shoulson & Fahn, 1979).

Exclusion criteria included use of anticonvulsant medications or benzodiazepines; commencement or change in dose of other psychotropic medications (i.e., anti-depressants, anti-psychotics) during the four weeks prior to, and during participation; choreiform movements precluding EEG data collection; a current episode of psychiatric illness, or a current substance use or alcohol use disorder as assessed and defined by the Mini International Neuropsychiatric Interview (MINI) 7.0.2 (Sheehan et al., 1997); and a history of significant head injury or traumatic brain injury, as defined by a loss of consciousness greater than 30 minutes or requiring a hospital admission.

Eleven of the 22 participants with HD had been formally diagnosed as having ‘manifest’ HD. Although the late premanifest and early manifest groups differed significantly with regards to average age, processing speed, TFC, and total motor symptoms, they did not differ significantly in average DBS scores (Manifest DBS M(SD) = 370.7(30.3), Range = 283.5-495.0; Late premanifest DBS M(SD) = 351.0(60.3), Range = 286.0-484.5, *t*_*20*_ *= −0*.*77, p = 0*.*45*). Nor did the groups differ significantly on the measures of apathy, anxiety, depression, general cognition, years of education, or whether they used psychotropic medications (refer to Table s1 in the supplementary materials for details of all comparisons). The highest total motor score for the combined sample was 49 out of a maximum score of 131, and therefore relatively mild. The similarity between the groups is consistent with the growing recognition of the non-motor onset and cognitive-psychiatric phenotypes in HD, thereby supporting our decision to use a combined sample (Paulsen et al., 2017, Ross et al., 2019). Fourteen participants in the combined HD group were taking stable doses of psychotropic medications comprising SSRIs (n = 8), risperidone (n=7), mirtazapine (n = 3), SNRIs (n = 2), and tetrabenazine (n = 1). One of the control participants was on a low dose tricyclic antidepressant as a migraine prophylactic.

Please refer to Table 1 for further demographic and clinical information for both the HD and control groups.

**Table 1.**
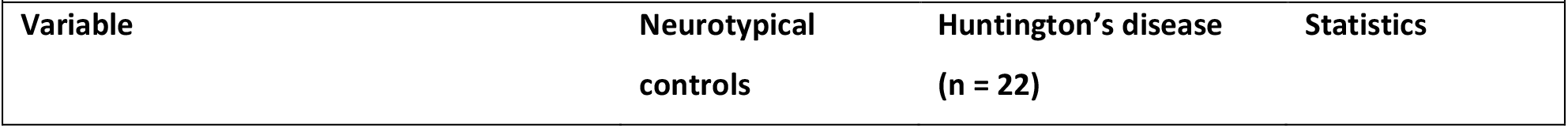
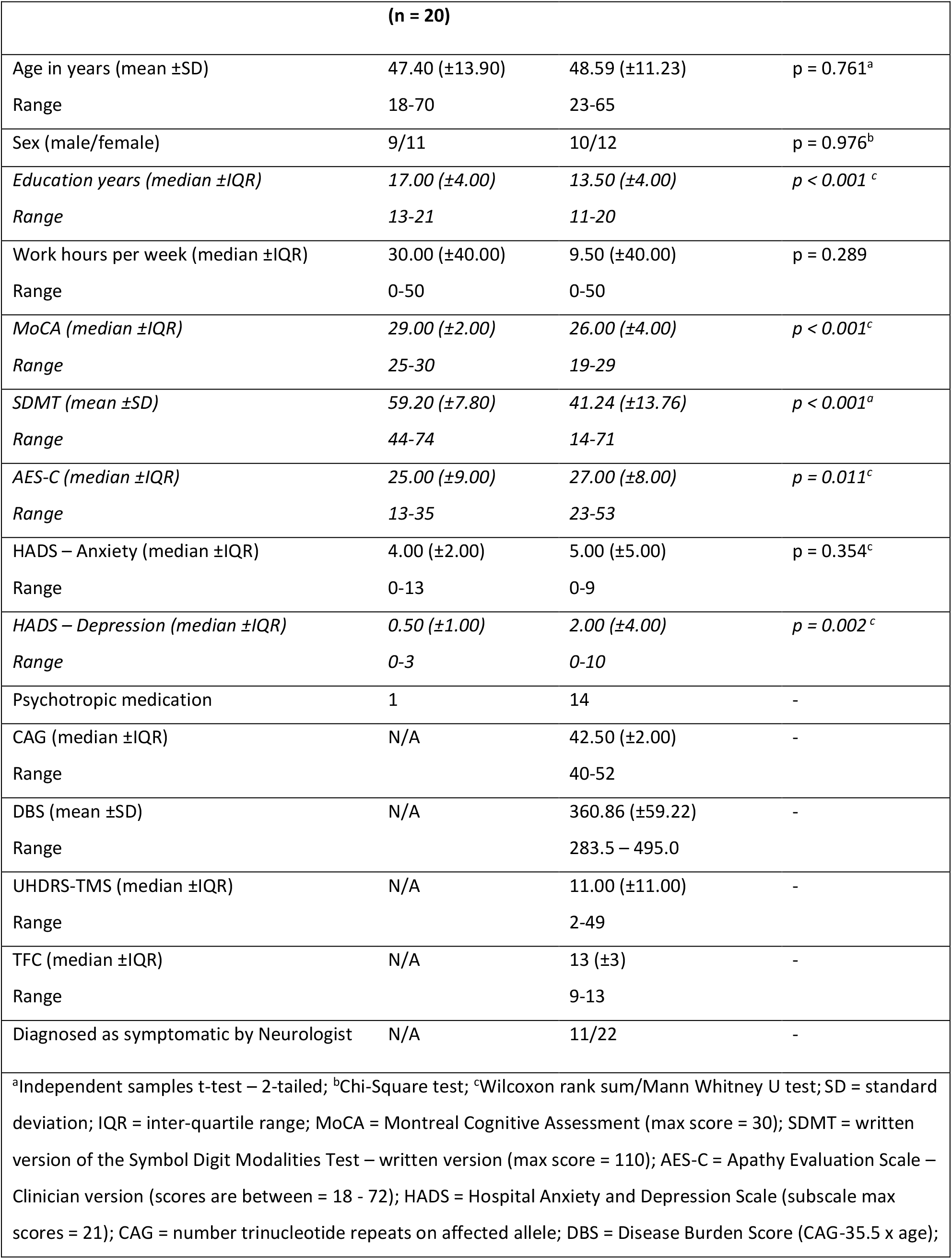
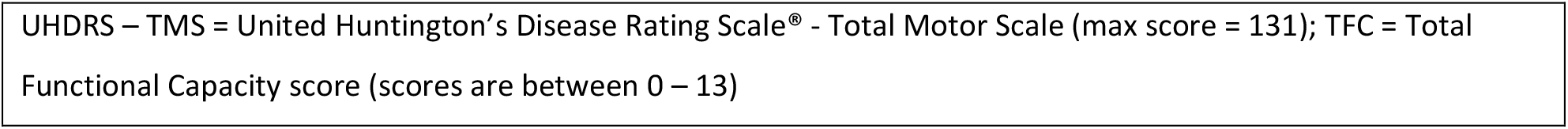
Characteristics of participant groups

### 2.3 Sample characterisation measures

The UHDRS® total motor scale (Kieburtz, 1996), Montreal Cognitive Assessment (MoCA) (Nasreddine et al., 2005), Hospital Anxiety and Depression Scale (HADS) (Zigmond & Snaith, 1983), Apathy Evaluation Scale – clinician version (Marin, Biedrzycki, & Firinciogullari, 1991), and Symbol Digit Modalities Test – written version (SMDT) (Smith, 1982) were administered as part of the study. The MoCA is a brief cognitive screen. The HADS was included to measure subclinical symptoms of anxiety and depression, with scores of ≤7 on both subscales considered to be within the ‘normal’ range (Snaith, 2003). The AES-C is a semi-structured clinical interview to assess the reductions in overt behaviour, emotional responsiveness and goal-directed thought content considered characteristic of apathy, with higher scores indicative of greater apathy (Marin et al., 1991). The SDMT is a timed, paper and pencil measure of processing speed that is sensitive, reliable, and well-validated for use in HD research (Stout et al., 2014).

All sample characterisation measures were administered by MCD, an experienced clinical neuropsychologist with certification in the administration of the UHDRS® total motor scale through the Enroll-HD clinical training portal (https://hdtraining.enroll-hd.org/). The UHDRS® total motor scale and MoCA data were not collected for three HD participants (two late premanifest and one manifest), and SDMT and AES-C data are missing for one HD participant due to the COVID-19 social distancing restrictions and lockdowns in Melbourne Victoria, which impacted data collection for a small number of participants.

### 2.4 EEG recording and data processing

Baseline resting state EEG was collected for three minutes with eyes closed, and three minutes with eyes open (fixating on a small black cross in the middle of a white screen) using a 45-channel montage via the Neuroscan EEG system with Ag/AgCl electrodes connected to a SynAmps amplifier (Compumedics, Australia). The electrodes used were: AF3, AF4, F7, F5, F3, F1, FZ, F2, F4, F6, F8, FC5, FC3, FC1, FCZ, FC2, FC4, FC6, T7, C3, C1, CZ, C2, C4, T8, CP5, CP3, CP1, CPZ, CP2, CP4, CP6, P7, P5, P3, P1, PZ, P2, P4, P6, P8, PO3, POZ, PO4, O1, OZ, O2. Recording was completed at a sampling rate of 1KHz, with impedances kept below 5kΩ, with the signal online referenced to CPz with the ground at FCz. The EEG data were pre-processed offline using the RELAX (Reduction of Electroencephalographic Artifacts) fully automated EEG pre-processing pipeline (Bailey, Biabani, et al., 2022; Bailey, Hill, et al., 2022) implemented in MATLAB (MathWorks, 2019) via the EEGLAB toolbox (Delorme & Makeig, 2004). Firstly, a notch filter (47<>52Hz) was applied to remove line noise, along with a zero-phase fourth order Butterworth filter (0.25<>80Hz) (Rogasch et al., 2017). The automated pipeline consisted of an initial extreme outlying channel and period rejection step, followed by a multi-channel Wiener filter algorithm which provided an initial reduction of eye movement, muscle activity and drift artifacts (Somers, Francart, & Bertrand, 2018). This was followed by subsequent reduction of remaining independent component analysis (ICA) artifacts identified using ICLabel via wavelet enhanced ICA (wICA) (Pion-Tonachini, Kreutz-Delgado, & Makeig, 2019). The initial bad electrode rejection was performed using the PREP pipeline (Bigdely-Shamlo, Mullen, Kothe, Su, & Robbins, 2015), and artifacts informing the Multiple Wiener filter algorithm were automatically identified using muscle artifact detection methods described by Fitzgibbon and colleagues (2016). After cleaning, the data were re-referenced to the average of all electrodes. The resting eyes closed data for one control participant had to be excluded due to an insufficient number of epochs remaining after pre-processing, while the resting eyes closed data for one HD participant was missing due to an equipment failure.

### 2.5 Spectral power

To separate periodic activity from the overall EEG signal, the data were processed using the extended Better OSCillation detection method (eBOSC) (Kosciessa et al., 2020). This required that the data be segmented into five-second epochs with 1.5 second overlaps on each side, leaving two second epochs for processing. The bilateral 1.5 second overlaps were removed to exclude edge artifacts associated with wavelet transformation (Kosciessa et al., 2020). The number of epochs available for analysis when segmented in this way were: Control M(SD) = 43.1(5.9) and HD M(SD) = 44.0(5.5). This method produced measures of periodic activity and 1/f activity (i.e., the background aperiodic component) from which a rhythmic signal-to-noise ratio (Rhythmic SNR) is derived for statistical analysis (Kosciessa et al., 2020). Specifically, Rhythmic SNR is “the background-normalized rhythmic amplitude as a proxy for the rhythmic 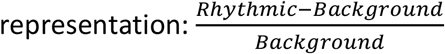” (Kosciessa et al., 2020, p. 6).

For comparison with previous research, group differences in spectral power of the combined periodic and aperiodic data (i.e., complete EEG signal) were computed in FieldTrip (Oostenveld, Fries, Maris, & Schoffelen, 2011) using a multi-taper fast Fourier transform and Hanning taper.

Power was averaged across all frequency bands between 1 and 45Hz and across all epochs. The results of these analyses of the complete EEG signal, please refer to the Supplementary Material.

### 2.6 Functional connectivity

For the functional connectivity EEG data comparisons, data were segmented into six-second epochs with 2.5 second overlaps on each side, which produced epochs with a unique one second period in the middle of each epoch, providing a buffer against the edge effects that can occur for connectivity analysis. This altered the number of epochs available for analysis to: Control M(SD) = 42.5(6.6) and HD M(SD) = 43.5(6.3) but was necessary to obtain epochs of sufficient length for robust connectivity estimates while protecting against both edge effects and the bias that occurs if connectivity values are extracted from overlapping data (Miljevic, Bailey, Vila-Rodriguez, Herring, & Fitzgerald, 2021). A surface Laplacian spatial filter was then applied to mitigate volume conduction (Miljevic et al., 2021). Lagged phase connectivity was calculated in FieldTrip using the debiased, weighted Phase Lag Index (wPLI) (Oostenveld et al., 2011; Vinck, Oostenveld, van Wingerden, Battaglia, & Pennartz, 2011). wPLI was calculated for all pairs of electrodes in all frequency bands between 1 and 45Hz as well as across epochs. ‘Lagged’ phase-based synchronisation analyses are preferred because they control for false positive findings resulting from volume conduction (i.e., simultaneous activation at multiple electrodes or regions of interest by the same underlying electrical field, which mimics synchronisation but in fact only reflects volume conduction of a single region or artifact) (Vinck et al., 2011).

### 2.7 Statistical analyses

Following removal of the aperiodic activity, between-group differences in periodic estimates of power within the delta (1-4Hz), theta (5-7Hz) and alpha (8-12Hz) bands were analysed separately in FieldTrip using non-parametric cluster-based permutation analyses (Maris & Oostenveld, 2007). A cluster was defined as being comprised of *at least* two adjacent electrodes with a p < 0.025. The Monte Carlo method was used to calculate the significance probability (2000 iterations). These analyses were also conducted on the combined periodic and aperiodic data, with the results presented in the Supplementary Material.

Between-group differences in functional connectivity within the delta, theta and alpha bands were analysed separately using Network Based Statistics (NBS) (Zalesky, Fornito, & Bullmore, 2010). Each of the 45 electrodes were defined as a ‘node’, and the strength of connectivity between two nodes (i.e., pair of electrodes) was defined as an ‘edge’. NBS uses a cluster-based permutation analyses approach, in that it utilises non-parametric permutation testing to control for multiple comparisons while maximising statistical power (Zalesky et al., 2010).

Significant connections between nodes are first identified based on a primary threshold, in this case we used the corresponding critical t-value for p < 0.025. Connections surpassing this threshold then undergo permutation testing (5000 permutations) with a family-wise error corrected significance alpha level; in this instance, with a secondary threshold of 0.05 (Zalesky et al., 2010). This second stage identifies ‘topological clusters’, in which nodes within a specific area are interconnected (Zalesky et al., 2010). Significant topological clusters were then visualised using the BrainNet viewer toolbox (Xia, Wang, & He, 2013).

Topological clusters and wPLI values that differed significantly between groups were extracted for further statistical analysis. Specifically, the average spectral power values from all electrodes within significant topological clusters were averaged together, and wPLI values for significant ‘edges’ identified in the connectivity analysis were averaged together and extracted for exploratory correlation with the AES-C and SDMT.

## 3.1 Results

### 3.2 Group differences in clinical measures

As depicted in Table 1, the HD group’s scores on the MoCA and SDMT were significantly lower than that of the control group (MoCA U_39_ = 45.50, p<.001; SDMT t_39_ = 5.10, *p<*.*001*). The control group also had a significantly higher number of years of education than the HD group (U_42_ = 77.50, *p<*.*001*). The HD group’s responses on the AES-C and the HADS depression subscale were significantly higher than those of the control group (AES-C U_41_ = 306.50, *p = .011*; HADS depression subscale U_42_ = 339.50, *p = .002*). Two participants with HD scored 8 and 10 respectively on the HADS depression subscale, putting them in the ‘mild’ (8-10) range for depression. Similarly, three neurotypical controls scored 8, 11 and 13 and one participant with HD scored 9 on the anxiety subscale, putting these participants in the ‘mild’ (8-10) and ‘moderate’ (11-14) ranges for anxiety on this screening measure (Stern, 2014). Note however, that no participants met criteria for a current psychiatric illness as assessed and defined by MINI.

### 3.3 Group differences in spectral power after removal of the aperiodic signal

#### 3.3.1 Resting eyes closed

Cluster-based permutation analysis of the periodic data (Rhythmic SNR) for all electrodes during the resting eyes closed condition found one diffuse significant *negative* cluster within the theta band (5-7Hz) (*p = .0004*) (Figure 1). This broad cluster of lower power in the HD group encompassed bilateral frontal, temporal, and occipital as well as left parietal electrodes. No significant between group differences were identified for power within the delta or alpha frequency bands (both *p > 0*.*05*). These findings accord with the results of the same analyses conducted on the complete EEG signal (i.e., periodic and aperiodic activity combined) (Supplementary Material Figure s1).

**Figure 1.**
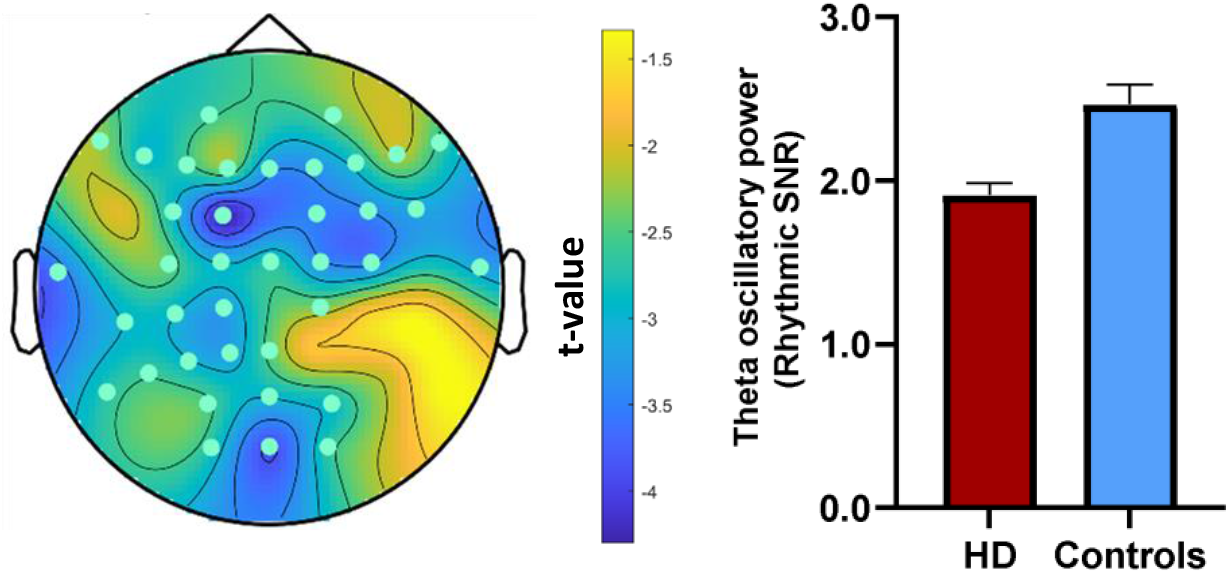
Left: Topographical plot of the large significant negative cluster of electrodes, indicating *lower* power in the theta band (5-7Hz) in the HD group relative to the control group using periodic data (Rhythmic SNR). Right: Graphical representation of group differences in average theta oscillatory power across significant electrodes (error bars reflect standard error).

#### 3.3.2 Resting eyes open

Cluster-based permutation analysis of the periodic data for all electrodes during the resting eyes open condition showed one significant *positive* cluster (increased power in the HD group) in bilateral frontocentral electrodes within the delta band (1-4Hz) (*p = .002*) (refer to Figure 2). There were no significant differences between groups for power within the theta or alpha frequency bands.

**Figure 2.**
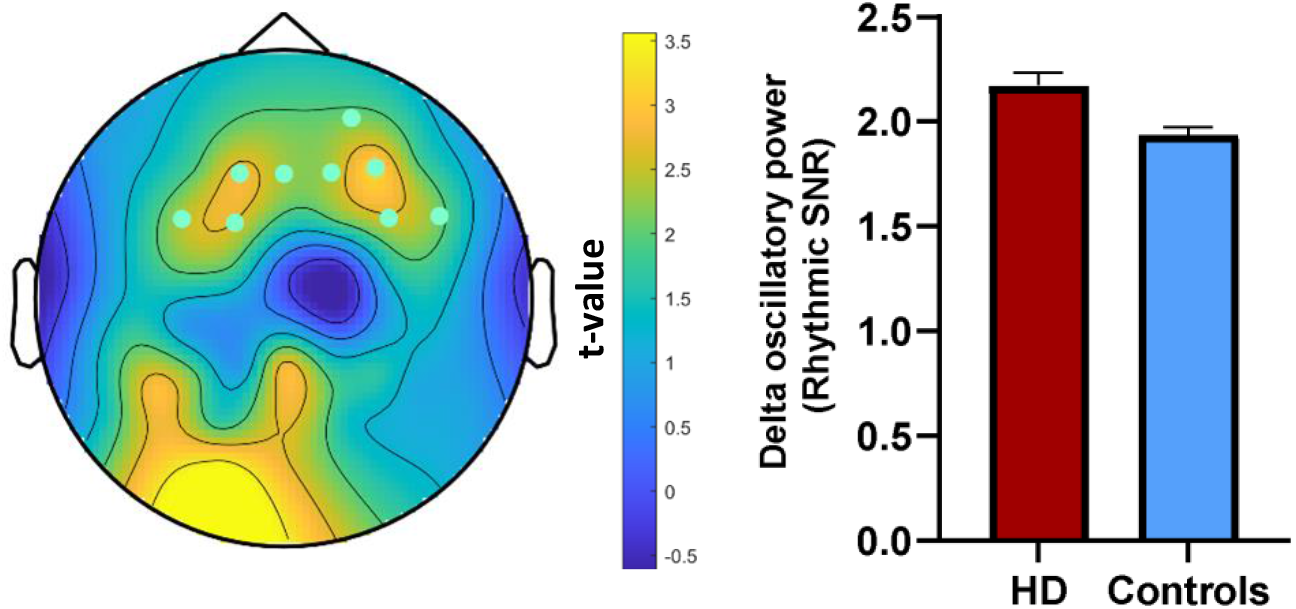
Left: Topographical plot of the positive cluster of frontocentral electrodes, suggesting *higher* power in the delta band (1-4Hz) in the HD group relative to the control group using periodic data only (Rhythmic SNR). Right: Graphical representation of group differences in average delta oscillatory power across significant electrodes (error bars reflect standard error).

These findings contrast with the results of the same analyses conducted on the complete EEG signal (including both periodic and aperiodic power), which found lower power within the theta and alpha bands, and no differences within the delta band in the HD group (Supplementary Material Figures s2-s3). Visual inspection of the spectral power distribution of the complete EEG signal (refer to Figure s4) suggests that differences in theta and alpha power detected using the complete EEG signal may reflect a broadband shift in the aperiodic 1/f activity, rather than actual differences in oscillatory power (Donoghue et al., 2020). Nevertheless, cluster-based permutation analysis of group differences in aperiodic activity (i.e., the intercept and slope of spectral distributions) did not reach significance (both *p >*.*05*).

### 3.4 Group differences in functional connectivity

The NBS analysis identified one significant diffuse network of *increased* functional connectivity within the delta band in the HD group during the resting eyes open condition, comprised of 40 nodes (electrodes) and 76 edges (connections between electrodes) (secondary threshold p = 0.04) (refer to Figure 3). There were no significant between-group differences in functional connectivity for the resting eyes closed condition.

**Figure 3.**
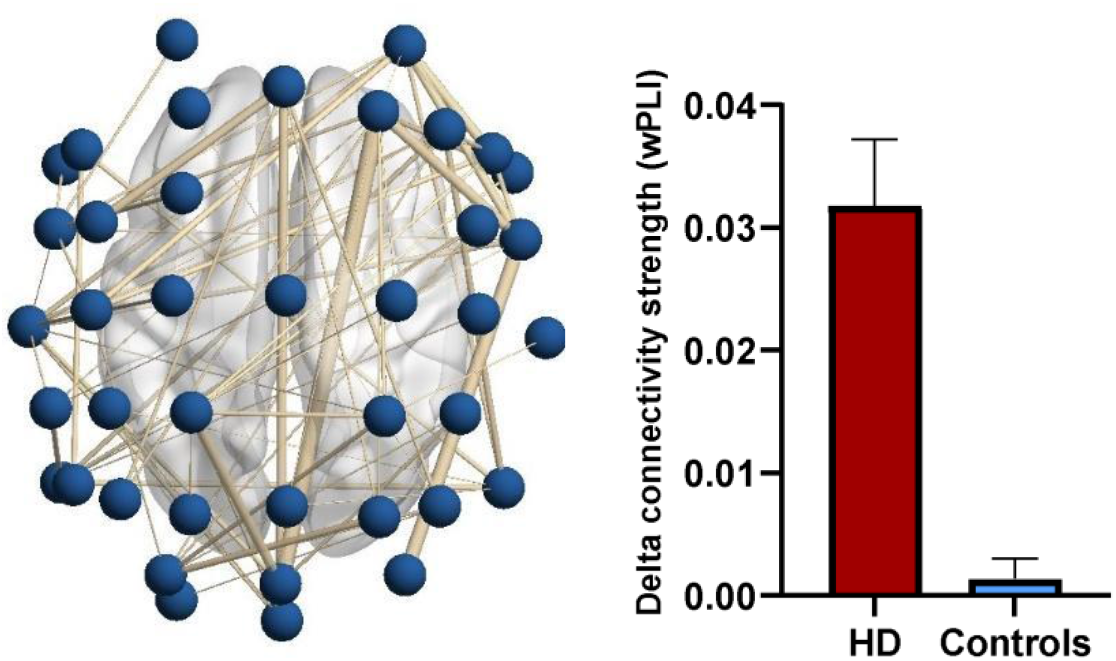
Left: Dorsal view of the network demonstrating significantly higher connectivity during the resting eyes open condition within the delta band (1-4Hz) in the HD group relative to the control group. Right: Graphical representation of delta connectivity strength taken from all edges comprising the significant subnetwork (error bars reflect standard error).

### 3.5 Correlations between neurophysiological markers, apathy, and processing speed

To limit the overall number of comparisons, only neurophysiological markers that significantly differed between the groups (i.e., theta oscillatory power; delta oscillatory power and delta connectivity strength) were assessed for correlations with the AES-C and SDMT. Given the exploratory goals of the correlations, no further adjustments were made for multiple comparisons (Greenland, 2021). There was one extreme outlier (i.e., a Z scores >±3.0) on the AES-C in the HD group, which was winsorized (i.e., 90^th^ percentile method) prior to further analysis. Non-parametric Spearman’s correlations were conducted to accommodate non-normally distributed variables where required. The results for each group are presented in Table 2.

**Table 2.**
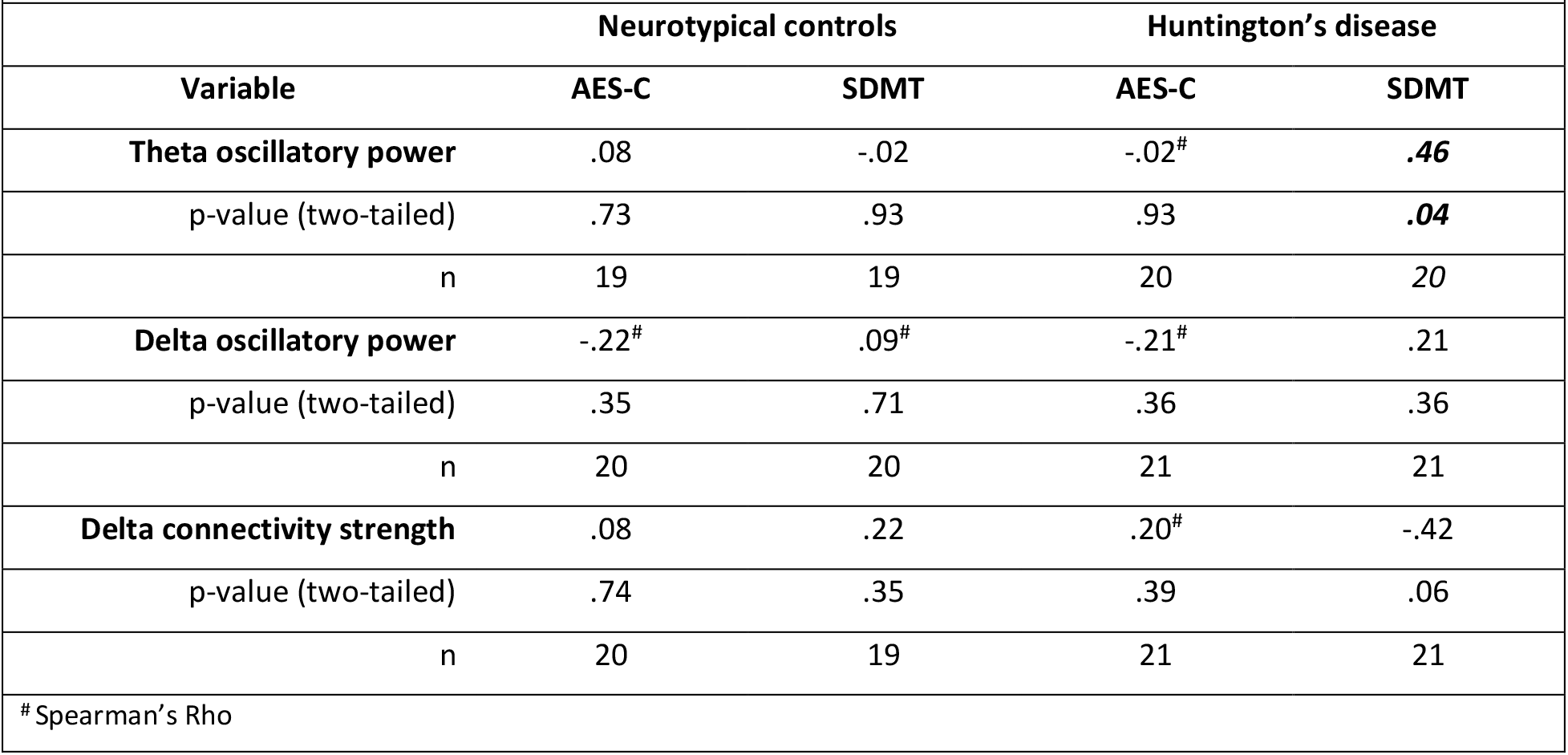
Correlation matrix between the significant neurophysiological markers, AES-C and SDMT within each group

For the HD group, the only statistically significant relationship was between theta oscillatory power and performance on the SDMT, *r* = .46, *n* = 20, *p = .04*, with higher theta oscillatory power during the resting eyes closed condition associated with higher SDMT scores. There were no significant correlations between any of the variables within the neurotypical control group. Selected scatterplots and additional correlation analyses between the neurophysiological markers and all sample characterisation measures for the HD group are presented in Table s2, and Figures s5 and s6 of the Supplementary Material. Lastly, there were no significant correlations between the neurophysiological metrics, HADS depression subscales scores, or years of education in either group (refer to Tables s3 and s4 of the Supplementary Material).

## 4.1 Discussion

We examined spectral power and lagged phase synchronisation in people with late premanifest and early-stage manifest HD and neurotypical controls. We found lower theta power in the HD group during the resting eyes closed condition in the periodic data; higher delta power in the HD group during the resting eyes open condition in the periodic data; higher connectivity within the delta band in the HD group during the resting eyes open condition; and a positive correlation between the HD group’s processing speed performance and theta power in the periodic data.

### 4.2 Resting eyes closed

In the resting eyes closed condition, our HD group had significantly lower oscillatory power within the theta band (5-7Hz) relative to neurotypical controls across bilateral frontal, temporal, occipital and left parietal electrodes. This finding was replicated using traditional measures of the EEG signal that did not account for non-oscillatory aperiodic 1/f activity (Supplementary Material). We did not, however, replicate previous findings of higher delta power and lower alpha power in the HD group relative to controls using either the periodic or complete EEG signal (with both periodic and aperiodic activity included).

Lower theta power in the resting eyes closed condition for the HD group is consistent with three previous studies (Odish et al., 2018; Painold et al., 2011; Ponomareva et al., 2014), but inconsistent with a number of others that found no group differences or *higher* theta power in the HD group (De Tommaso et al., 2003; Delussi et al., 2020; Hunter et al., 2010; Painold et al., 2010; Piano, Imperatori, et al., 2017; Piano, Mazzucchi, et al., 2017; Van Der Hiele et al., 2007). Nevertheless, our findings were consistent across analyses of both the periodic and complete EEG signal.

Inconsistency within the HD literature regarding theta power may be due to inconsistent definitions of the theta band, which vary within the range of 3.5 to 8Hz. Like our study, two of the three previous studies that found lower theta activity in HD relative to neurotypical controls did not include frequencies below 5Hz within their definition of theta (the third did not specify bandwidth limits), an approach which may have avoided conflation of delta and theta activity (Painold et al., 2011; Ponomareva et al., 2014). There is similar variability in the definitions of delta and alpha bands (i.e., definitions for delta have specified various cut-off points between 0.5 and 6Hz, and definitions for alpha have specified various cut-off points between 8 and 13.5Hz), but significant between group differences for these frequencies are more consistent across studies. The present study is otherwise methodologically consistent with most previous investigations of resting state EEG in HD, so the reason for our failure to find higher delta power and lower alpha power in the resting eyes closed condition when analysing either the periodic or the complete EEG signal may reflect the inherent variability in the electrophysiological manifestation of HD.

### 4.3 Resting eyes open

In the resting eyes open condition, our HD group had significantly higher oscillatory power in frontocentral electrodes in the delta band (1-4Hz) than that of the neurotypical control group after controlling for aperiodic activity. By contrast, analyses of the complete EEG signal showed lower power in the theta and alpha bands, and no significant between-group differences in the delta band (Supplementary Material). Previous studies have not collected resting state data with participants’ eyes open, limiting the comparability of the current findings with previous research (Barry & De Blasio, 2017; Boytsova & Danko, 2010). Higher delta power within the complete EEG signal during the resting eyes closed state, however, has been a consistent finding in previous HD research (Delussi et al., 2020; Leuchter et al., 2017; Nguyen, Bradshaw, Stout, Croft, & Georgiou-Karistianis, 2010; Odish et al., 2018).

Increased delta power in resting state EEG recordings has been associated with a variety of neurodegenerative brain disorders (Knyazev, 2012), as well as in response to neural plasticity induction (Assenza, Pellegrino, Tombini, Di Pino, & Di Lazzaro, 2015), and recovery post-stroke (Cassidy et al., 2020). Based primarily on animal studies, the generation of delta oscillatory activity in the human brain has also been attributed to nuclei within the cortico-basal ganglia-thalamocortical networks (Knyazev, 2007; Zang, 2009), which are preferentially affected by HD pathology (Waldvogel et al., 2015). As such, our finding of higher delta power concentrated around frontocentral electrodes in the HD group may be the oscillatory reflection of ongoing degenerative and compensatory processes (e.g., Soloveva et al., 2020).

The potential for the combined periodic and aperiodic signal to result in spurious power estimates may explain our contrasting periodic and complete EEG signal findings in the resting eyes open condition (Kosciessa et al., 2020). Visual comparison of the spectral power distributions for the complete EEG signal during the resting eyes open condition (Supplementary Material Figure s4) suggests group differences in aperiodic activity caused the overall power distribution of neurotypical controls to be higher than the overall distribution of the HD group (a so called ‘broadband shift’). This broadband shift may be responsible for the lower theta and alpha power in the HD group in that analysis, rather than the differences reflecting true differences in oscillatory power (Donoghue et al., 2020). This is speculative, however, given that the differences in aperiodic activity between the groups were not statistically significant when measured separately from the periodic activity. A recent EEG-fMRI study of neurotypical controls resting with eyes open found that *lower* aperiodic activity correlated with *higher* hemodynamic activity in prefrontal regions (Jacob, Roach, Sargent, Mathalon, & Ford, 2021). It may be that lower aperiodic activity in the HD group during the eyes open condition reflected greater activation of prefrontal cognitive control networks to sustain attentional control and compensate for the impact of HD pathology on frontostriatal networks (Aracil-Bolanos et al., 2022; Jacob et al., 2021; Pini et al., 2020). These group differences may also only have emerged in the eyes open condition due to the increased need for cognitive control precipitated by the external demands on attention that accompany eyes open paradigms (Barry & De Blasio, 2017; Boytsova & Danko, 2010).

### 4.4 Functional connectivity

Our HD group had significantly higher strength of lagged phase connectivity in the delta band in the eyes open condition; a finding that is partially consistent with the two previous studies of EEG functional connectivity in HD, both of which implemented an eyes closed procedure (Delussi et al., 2020; Piano, Imperatori, et al., 2017). Within the context of neurodegenerative diseases, higher delta connectivity appears to be associated with diseases preferentially affecting the frontal networks. For example, relative to neurotypical controls, delta connectivity has been found to be similar or lower in people with Alzheimer’s dementia (Hata et al., 2016; Yu et al., 2016), but higher in people with Parkinson’s disease (Sanchez-Dinorin et al., 2021), Amyotrophic Lateral Sclerosis (Iyer et al., 2015), and behavioural variant Frontotemporal Dementia (Yu et al., 2016). We speculate that, like higher delta oscillatory power, higher delta connectivity may be an electrophysiological assay of ongoing, frontally concentrated degenerative and compensatory processes.

### 4.5 Correlations

We found no significant correlations between our measure of apathy (AES-C) and the neurophysiological markers found to differentiate the HD group from the neurotypical control group. We suspect this was in large part due to insufficient spread of scores on the measure of apathy (refer to Figure s5 in Supplementary Material) and clinical apathy not being an inclusion criterion for participants (the latter would have made recruitment unfeasible for such a resource intensive study of a rare disease). Sensitive and specific measures of apathy across all stages of HD remains an ongoing challenge (Atkins, Andrews, Chong, & Stout, 2021; Atkins et al., 2020). Of the other correlations conducted, only the association between processing speed (SDMT) and eyes closed theta oscillatory power in the HD group was statistically significant, indicating that HD participants with higher theta oscillatory power demonstrated better performances on the SDMT. To our knowledge, this is the first study to find an association between theta power in the periodic or complete EEG signal and SDMT performance in people with HD. Direct investigations of the relationship between SDMT performance and resting theta power are lacking in the broader literature, but a positive association between oscillations in the theta (and other) frequency bands and higher performance in a variety of cognitive processes has been well-documented (e.g., Karakas, 2020; Klimesch, 1999).

### 4.6 Relevance to future research

This study extends previous research into the neurophysiology of HD by examining resting eyes closed as well as resting eyes open EEG, and the strength of oscillatory power following removal of aperiodic activity. As our understanding of the functional significance of periodic and aperiodic activity increases along with our ability to modulate these variables through treatments (Başar & Güntekin, 2013; Klink, Passmann, Kasten, & Peter, 2020; Schutter & Wischnewski, 2016), so too does our need for separate data processing and analyses of periodic and aperiodic activity (Donoghue et al., 2020; Kosciessa et al., 2020). This study demonstrates the removal of aperiodic activity from the EEG signal using the eBOSC method (Kosciessa et al., 2020) and how the analysis of the isolated periodic signal may refine our understanding of oscillatory activity in HD. This is vital, as finding effective treatments for non-motor symptoms of HD symptoms remains a priority and requires the development of sensitive, specific, non-invasive, and inexpensive proximate markers which might indicate which neurophysiological aspects treatments should target (Ferreira et al., 2022; Stout et al., 2017).

Evidence for the reliability of quantitative EEG as a sensitive analogue of clinical symptoms in neurodegenerative diseases has significantly increased over the last decade (Geraedts et al., 2018; Sanchez-Reyes, Rodriguez-Resendiz, Avecilla-Ramirez, Garcia-Gomar, & Robles-Ocampo, 2021; Shim & Shin, 2020). The neurophysiological markers generated by EEG are also sensitive to different treatment modalities (e.g., non-invasive brain stimulation, pharmacological agents, behavioural therapies) (Ahn, Prim, Alexander, McCulloch, & Fröhlich, 2018; Best, Gale, Tran, Haque, & Bowie, 2019; Hua et al., 2022). This study adds to a small, but relatively consistent body of literature on the potential utility of quantitative EEG as a proximate marker for non-motor symptoms in HD.

### 4.7 Limitations

Our sample size was comparable to other studies of resting state EEG in HD (De Tommaso et al., 2003; Odish et al., 2018; Piano, Imperatori, et al., 2017; Piano, Mazzucchi, et al., 2017; Van Der Hiele et al., 2007). Nevertheless, a larger sample size of people with HD would have been preferable to increase statistical power and generalizability of the findings. While we intentionally contained our sample to the mildest end of the disease spectrum, broadening our sample to include people in later stages may have revealed additional neurophysiological findings. Extending our sample in this manner, however, was beyond the scope of this study. As previously noted, our choice of apathy measure lacked the sensitivity required to detect variation within our groups, thereby affecting our correlation analyses. The difference between the groups in years of education was also a limitation.

### 4.8 Conclusion

Effective treatments for non-motor symptoms of HD remain a priority for the HD community. Clinical trials for non-motor symptoms require sensitive, non-invasive, and inexpensive proximate markers. Our findings add to the existing literature on the utility of quantitative spectral power and functional connectivity metrics generated from short periods of resting state EEG data as sensitive and objective symptom markers for clinical research in HD.

## Data Availability

All data produced in the present study are available upon reasonable request to the authors from genuine researchers, provided they agree to preserve the confidentiality of the information.

## Acknowledgements

MCD was supported by the Research Training Program Stipend and a Monash Graduate Excellence Scholarship. KEH was supported by National Health and Medical Research Council (NHMRC) Fellowships (1082894 and 1135558). PBF was supported by an Investigator Fellowship from the NHMRC (1193596). ATH was supported by an Alfred Deakin Postdoctoral Research Fellowship. We gratefully acknowledge the time, involvement, and feedback from the participants. This research would not have been possible without them. We also acknowledge the assistance with participant recruitment provided by the Statewide Progressive Neurological Disease Service at Calvary Health Care Bethlehem.

## Disclosures

KEH is a founder of Resonance Therapeutics. PBF has received equipment for research from MagVenture A/S, Nexstim, Neuronetics and Brainsway Ltd and funding for research from Neuronetics. PBF is a founder of TMS Clinics Australia and Resonance Therapeutics. JCS is founder and a director of Zindametrix which provides cognitive assessment services in HD clinical trials, and Stout Neuropsych, which provides consultancy services for pharmaceutical companies. MCD, ATH, and NWB have no biomedical financial interests or potential conflicts of interest to report.

## Supplementary Materials

**Table s1.** Comparison of baseline measures in HD participants according to diagnostic status

|  |  |  |  |  |
| --- | --- | --- | --- | --- |
| **Variable** | **Late premanifest** | **Early manifest** | **Statistics** |  |
| *Age in years (mean ±SD)* | 43.64 (±10.61) | 53.55 (±10.04) | $t_{20} = -2.25$ | $p = .04$ |
| *Range* | 23-63 | 30-65 |  |  |
| Education years (mean ±SD) | 14.36 (±2.38) | 13.91 (±2.60) | $t_{20} = 0.43$ | $p = .67$ |
| Range | 11-18 | 11-20 |  |  |
| MoCA (median ±IQR) | 26 (±4) | 25 (±4) | $U_{19} = 30.50$ | $p = .24$ |
| Range | 21-29 | 19-29 |  |  |
| *SDMT (mean ±SD)* | 47.45 (±13.63) | 34.40 (±10.75) | $t_{19} = 2.42$ | $p = .03$ |
| *Range* | 29-71 | 14-53 |  |  |
| AES-C (median ±IQR) | 27 (±5) | 29 (±9) | $U_{21} = 64.00$ | $p = .56$ |
| Range | 25-41 | 24-53 |  |  |
| HADS – Anxiety (median ±IQR) | 5 (±3) | 2.50 (±6) | $U_{22} = 51.50$ | $p = .56$ |
| Range | 1-8 | 0-9 |  |  |
| HADS – Depression (median ±IQR) | 1 (±2) | 3 (±6) | $U_{22} = 76.50$ | $p = .30$ |
| Range | 1-7 | 0-8 |  |  |
| *UHDRS-TMS (median ±IQR)* | 4 (±4) | 14 (±27) | $U_{19} = 81.50$ | $p = .001$ |
| *Range* | 2-16 | 7-49 |  |  |
| DBS (mean ±SD) | 351.05 (±60.32) | 370.70 (±59.30) | $t_{20} = 0.77$ | $p = .45$ |
| Range | 286-484.50 | 283.50-495 |  |  |
| *TFC (median ±IQR)* | 13 (±0) | 10 (±3) | $U_{22} = 18.50$ | $p = .004$ |
| *Range* | 12-13 | 9-13 |  |  |
| Medication all types | 5 | 9 | $X^2 = 3.14$ | $p = .08$ |
| <sup>a</sup>Independent samples t-test – 2-tailed; <sup>b</sup>Chi-Square test; <sup>c</sup>Wilcoxon rank sum/Mann Whitney U test; SD = standard deviation; IQR = inter-quartile range; MoCA = Montreal Cognitive Assessment (max score = 30); SDMT = written version of the Symbol Digit Modalities Test – written version (max score = 110); AES-C = Apathy Evaluation Scale – Clinician version (scores are between = 18 - 72); HADS = Hospital Anxiety and Depression Scale (subscale max scores = 21); DBS = Disease Burden Score (CAG-35.5 x age); UHDRS – TMS = United Huntington's Disease Rating Scale® - Total Motor Scale (max score = 131); TFC = Total Functional Capacity score (scores are between 0 – 13) |  |  |  |  |

### Group differences in EEG spectral power

The complete EEG signal (periodic and aperiodic activity) was segmented into non-overlapping three-second epochs (number of epochs remaining for analysis: Controls M(SD) = 51.4(5.8) and HD M(SD) = 53.2(5.7)).

### Resting eyes closed

Cluster-based permutation analysis comparing all electrodes during baseline resting eyes closed EEG found no significant differences between groups for power within the delta or alpha frequency bands (both *p > 0*.*05*). There was, however, one diffuse *negative* significant cluster identified within the theta band (5-7Hz) (*p = .002*) (refer to Figure s1).

**Figure s1.**
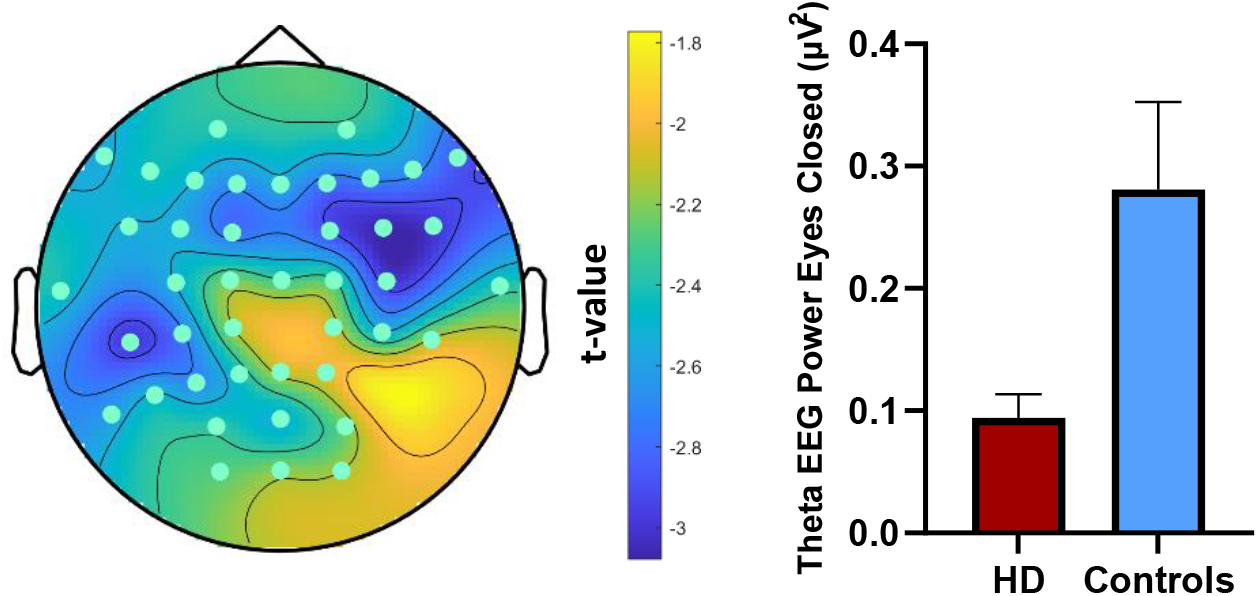
Left: Topographical plot of the large significant negative cluster of electrodes, suggesting *lower* power in the theta band (5-7Hz) during the resting eyes closed condition in the HD group relative to the control group. Right: Graphical representation of group differences in average theta EEG power across the significant electrodes (error bars reflect standard error).

### Resting eyes open

Cluster-based permutation analysis comparing all electrodes during resting eyes open EEG found no significant differences between groups for power within the delta frequency band. By contrast, there were significant *negative* clusters identified within the theta (*p = 0*.*011*) and alpha (*p = 0*.*025*) bands (refer to Figures s2 and s3).

**Figure s2.**
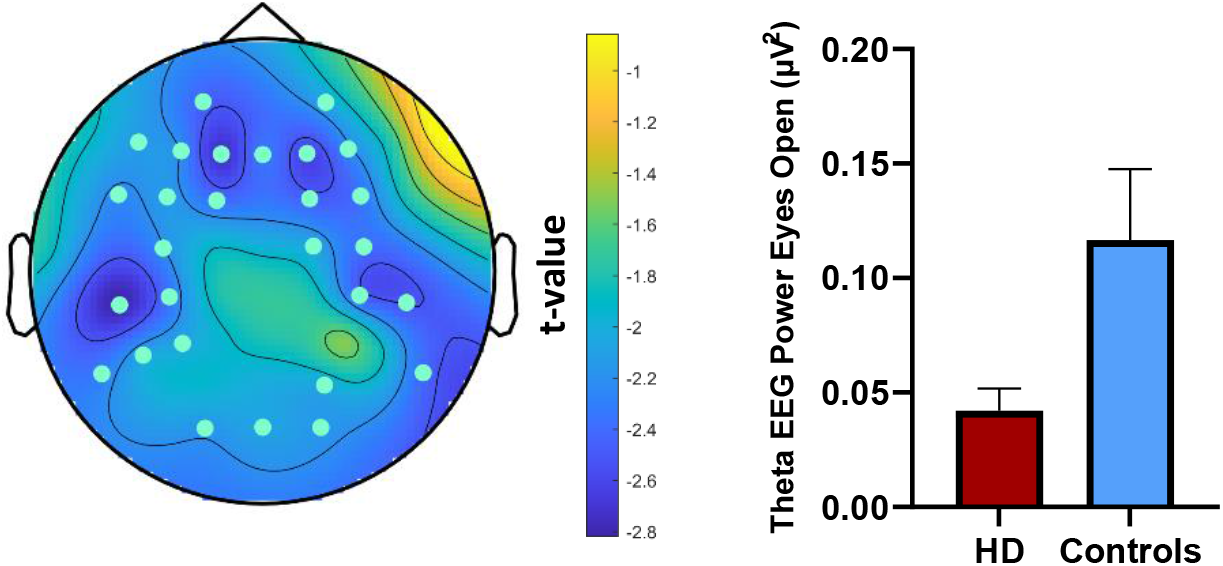
Left: Topographical plot of the large significant negative cluster of electrodes, suggesting *lower* power in the theta band (5-7Hz) during the resting eyes open condition in the HD group relative to the control group. Right: Graphical representation of group differences in average theta EEG power across significant electrodes (error bars reflect standard error).

**Figure s3.**
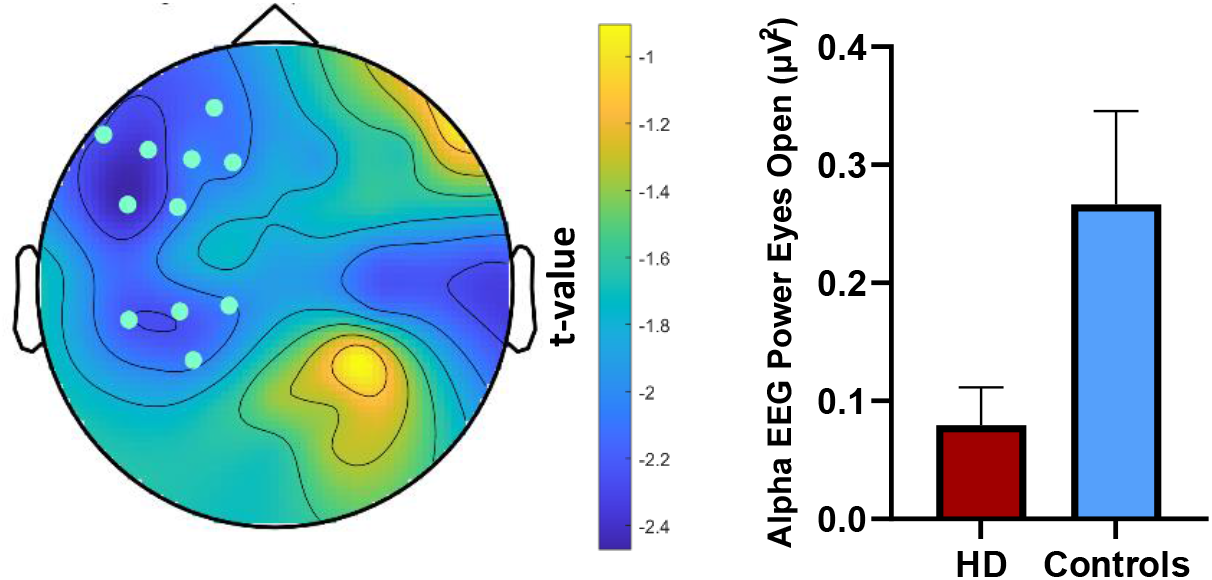
Left: Topographical plot of the significant negative cluster of left frontoparietal electrodes, suggesting *lower* power in this region specific to the alpha band (8-12Hz) during the resting eyes open condition in the HD group relative to the control group. Right: Graphical representation of group differences in average alpha EEG power across significant electrodes (error bars reflect standard error).

**Figure s4.**
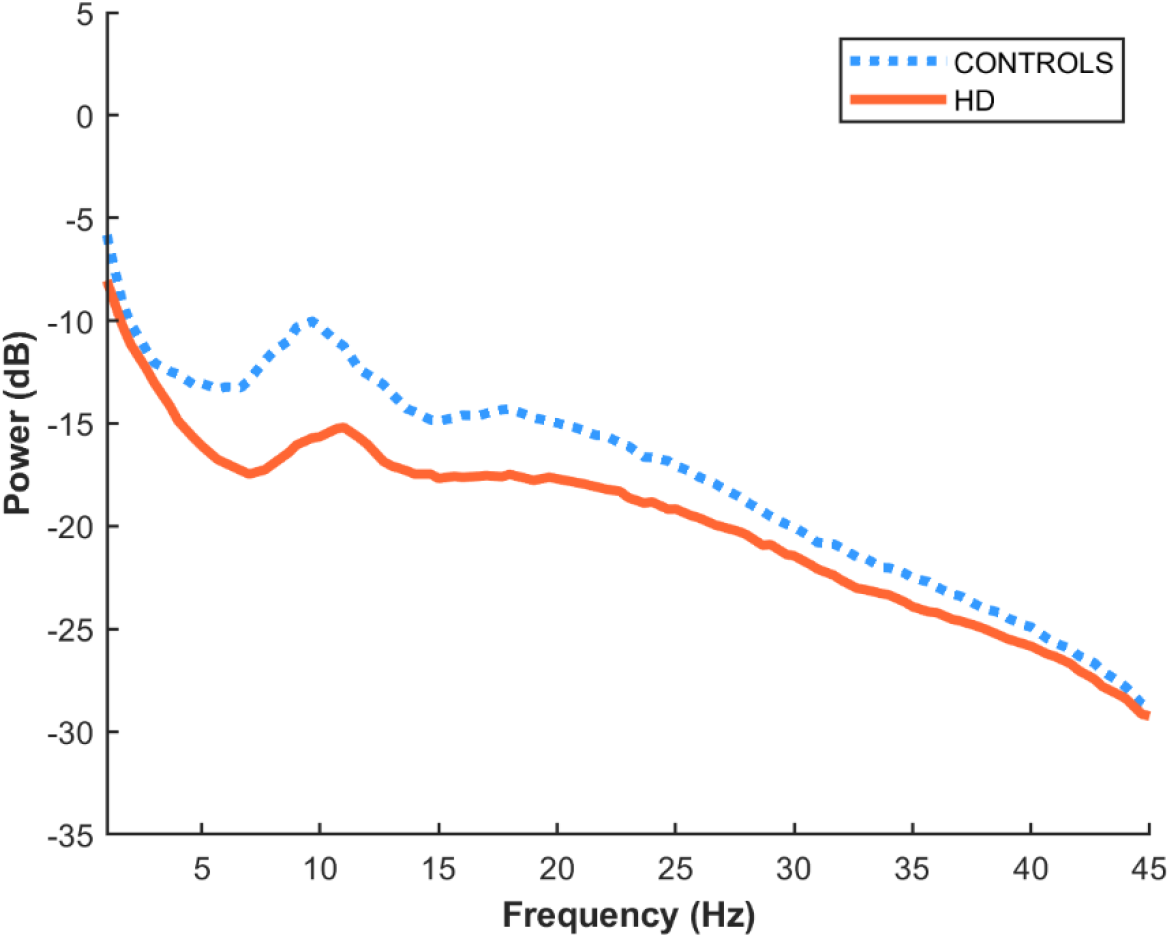
Spectral power distribution of each group averaged across all electrodes during resting state eyes open prior to removal of the aperiodic signal.

**Table s2.**
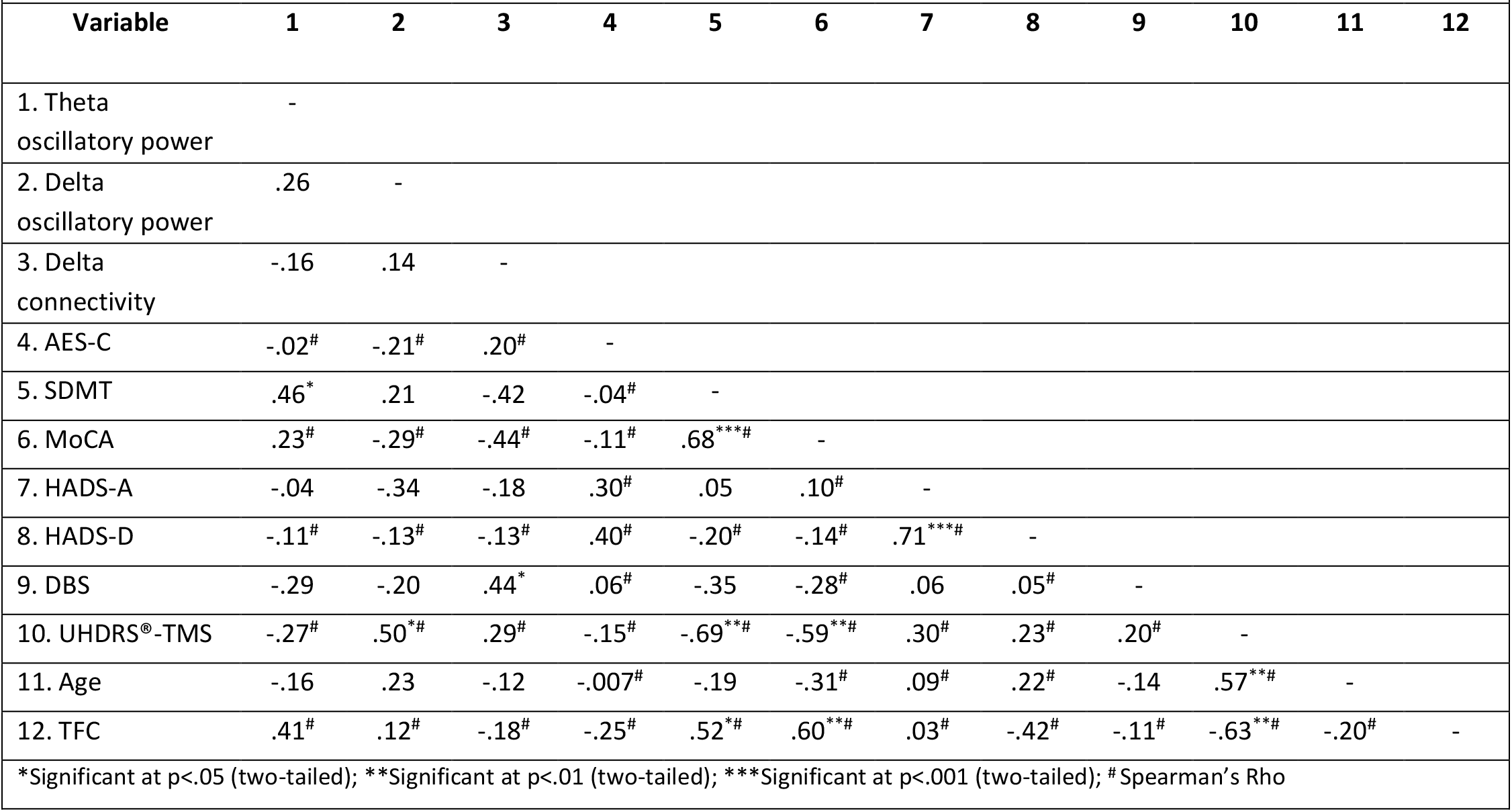
Correlation matrix between significant neurophysiological markers and all sample characterisation measures in the HD group

**Table s3.**
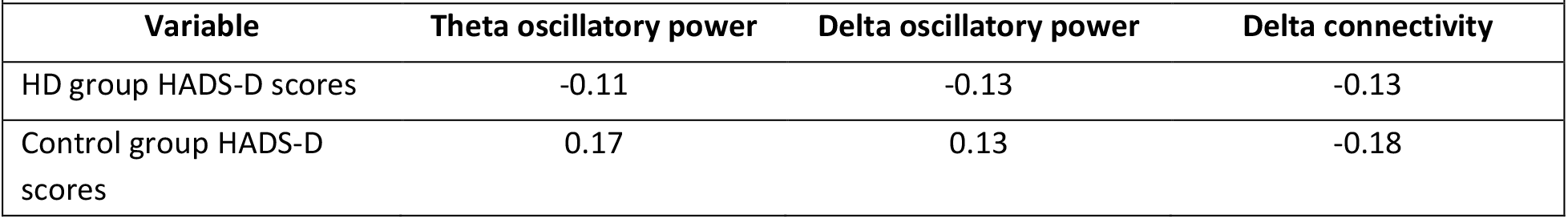
Spearman correlations between significant neurophysiological metrics and HADS-D scores for both groups

**Table s4.**
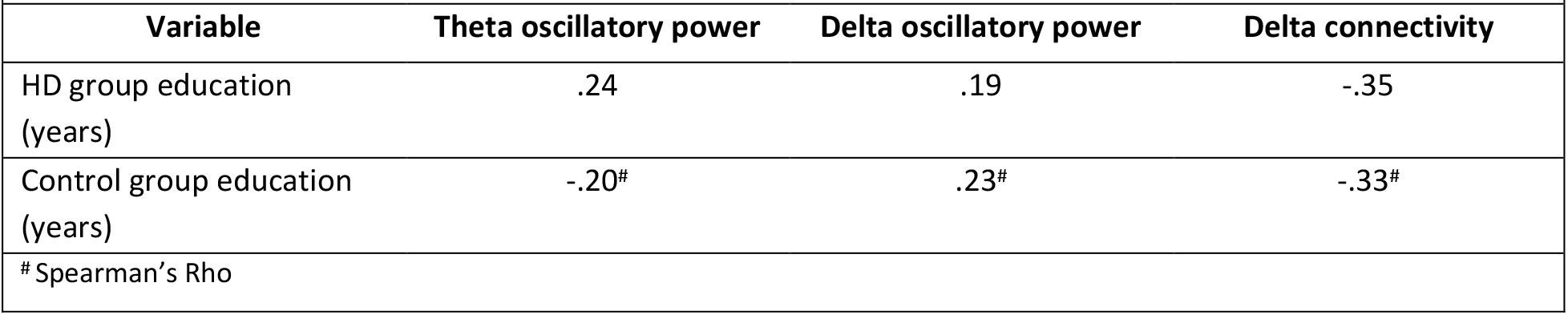
Correlations between significant neurophysiological metrics and years of education for both groups

| Variable | Theta oscillatory power | Delta oscillatory power | Delta connectivity |
| --- | --- | --- | --- |
| HD group education (years) | .24 | .19 | -.35 |
| Control group education (years) | -.20<sup>#</sup> | .23<sup>#</sup> | -.33<sup>#</sup> |
| # Spearman's Rho |  |  |  |

**Figure s5.**
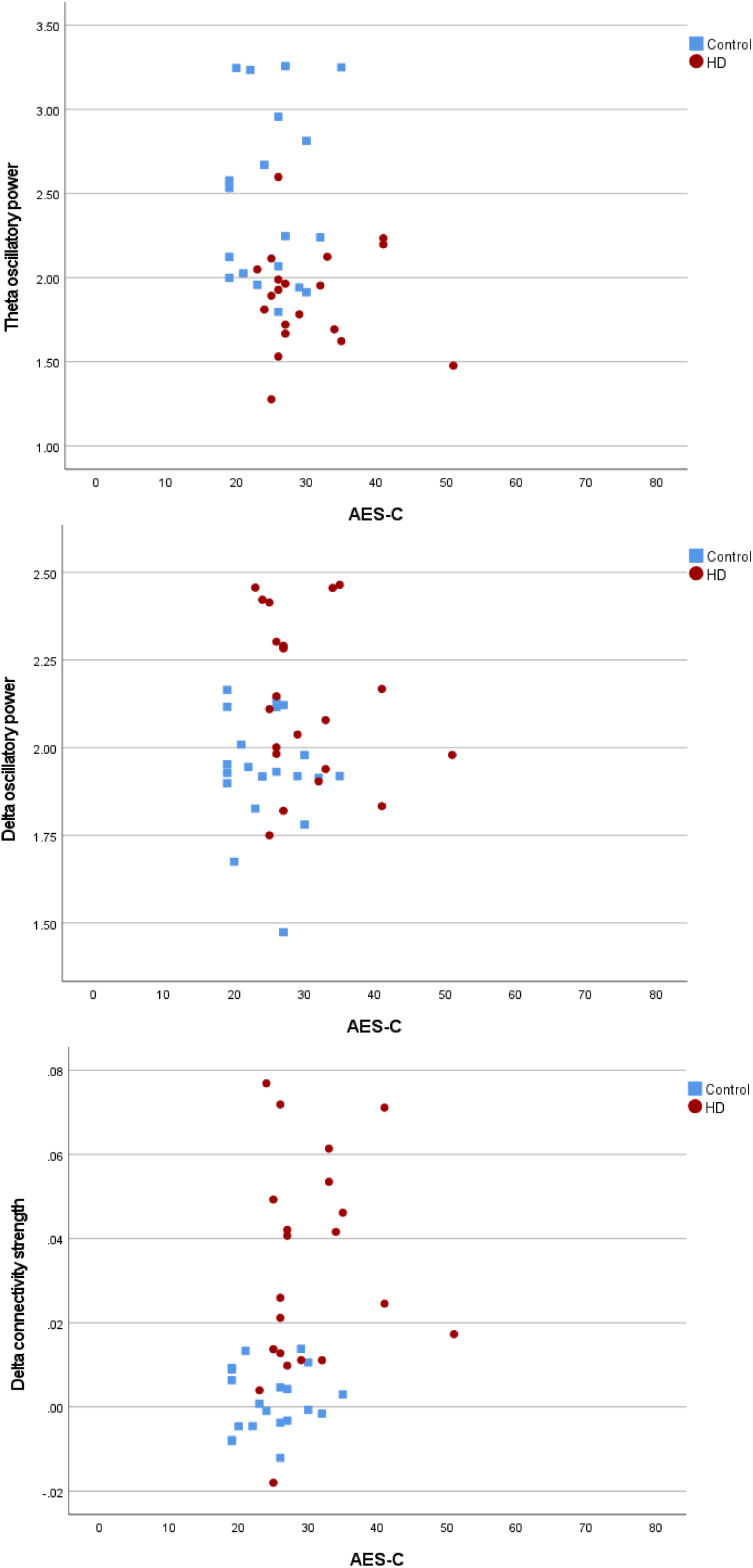
Scatterplots of the AES-C and significant neurophysiological markers.

**Figure s6.**
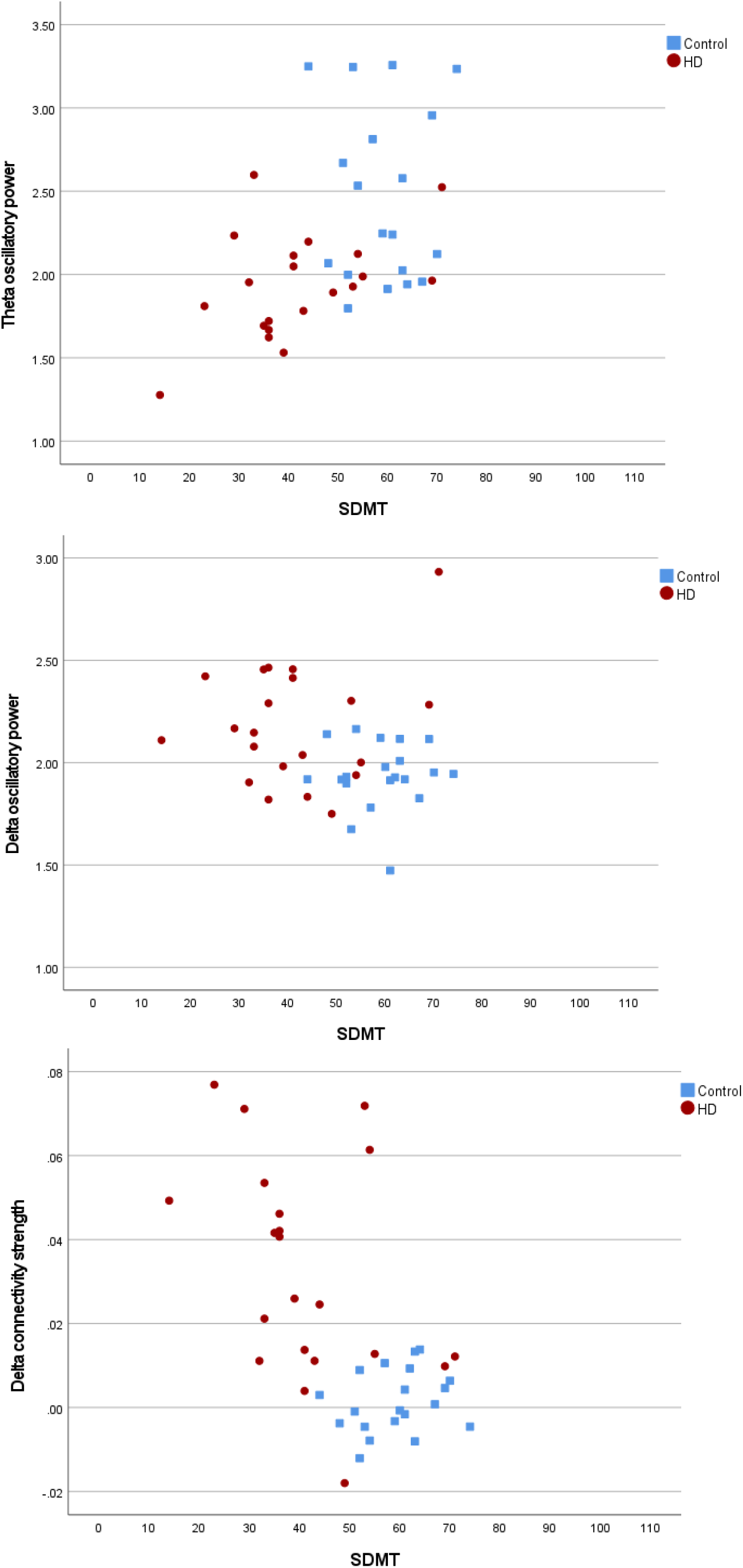
Scatterplots of SDMT and significant neurophysiological markers.

## Notes

### Clinical Trial

Australian New Zealand Clinical Trials Registry (ANZCTR): 12619000870156

### Author Declarations

Alfred Health Ethics Committee gave ethical approval for this work. Calvary Health Care Bethlehem Research Ethics and Ethics Committee gave ethical approval for this work.

### Summary of Updates

More detailed description of the sample of participants with HD.

